# Anxiety and Depression in Patients with Pulmonary Hypertension and its Impact on Health-Related Quality of Life

**DOI:** 10.1101/2021.04.22.20232645

**Authors:** Sarfraz Saleemi, Reem Alameer, Toka Alsulaim, Naif S Alghasab

## Abstract

**Background:** Pulmonary Hypertension (PH) is a chronic progressive disease which affects quality of life. The prevalence of mood disorders in PH patients in Saudi Arabia is not known.

**Objectives:** This is a prospective observational study to investigate the prevalence of anxiety and depression in patients with pulmonary hypertension and its effect on health-related quality of life (HRQoL).

**Methods:** Forty-two PH patients were evaluated for mood disorders by using Hospital Anxiety and Depression Scale (HADS) and health related quality of life was assessed with The Short Form 36 (SF-36) Health Survey questionnaire.

**Results:** Mean age for the cohort was 42.8 (±13.3) years and 31(73%) were female. Mean pulmonary artery pressure was 46.4 (± 15) mm Hg and 25 (60%) patients had severe PH (mPAP ≥45 mmHg). Thirty-eight (90%) patients were in World Health Organization Functional Class (WHO-FC) II/III and 5 (10%) class IV. HADS anxiety and depression score was positive (11-21) in 4 (10%) and 6 (14%) patients respectively. Presence of anxiety and/or depression affected HRQoL.

**Conclusion:** Prevalence of anxiety and depression in PH patients in Saudi Arabia is low (10% and 14% respectively) compared to published studies. The low prevalence may be explained by cultural and social differences and strong family support.

## Introduction

Pulmonary hypertension (PH) is a chronic progressive disease that carries high mortality and morbidity.^1^ Dyspnea on exertion due to elevated pulmonary artery pressure, increased pulmonary vascular resistance, and right heart failure, is the most common symptom which limits daily physical activity.^2^ The quality of life of patients with pulmonary hypertension is affected from many aspects, such as limitation of the physical activity that may lead to social isolation and unemployment. As a result, the patients are at risk of developing mental health problems such as depression and anxiety.^3,4^

Mental disorders such as depression and anxiety are common in PH patients. Löwe B et al. reported that depression was related to the degree of symptoms and functional limitation. The prevalence of major depression increased from 7.7% in patients with New York heart association (NYHA) functional class (FC) I to 45% in FC-IV.^5^ McCollister DH et al. looked into the prevalence of depression in patients with PH in the outpatient setting. The result of the study showed that up to 55% of PH patients seen in two PH referral centers in the United States suffered from depressive symptoms and 15% had major depressive disorder.^6^ Another study found that the prevalence of depression and anxiety in PH patients was 53% and 51% respectively.^7^

The Largest PH registry in US called REVEAL Registry (Registry to Evaluate Early and Long-term PAH Disease Management) has shown that 25% of PH patients has depression compared to 6.7% in general population.^8^ Other mood disorders are also common in these patients. An international survey of PH patients revealed the presence of feeling of worthlessness (22%), frustration (35%), anger (24%), and reduced pleasure in activities (25%) that they enjoyed prior to diagnosis of PH.^9^

King Faisal Specialist Hospital and Research Center has a dedicated pulmonary hypertension program. The prevalence of depression and anxiety in patients suffering from PH in the Arab world is never been reported where family and cultural traditions are different than in the western world. This provides a chance to study the prevalence of depression and anxiety in this group of patients and to look into their impact on the quality of life.

## Methods

### Study Design

This is a prospective, observational study which includes adult patients ≥14 years of age with a diagnosis of pulmonary hypertension (PH) confirmed by right heart catheterization and defined as mean pulmonary artery pressure (mPAP) ≥ 25 mmHg, pulmonary capillary wedge pressure (PCWP) < 15 and pulmonary vascular resistance (PVR) >3 wood units.^10^ Pulmonary hypertension was categorized into mild, moderate and severe based on mPAP (25-34, 35-44, ≥45 mmHg respectively).^11^ The patients were on PH-specific treatment for at least 3 months and had regular follow up at pulmonary hypertension clinic. Patients aged <14 year of age, pregnant ladies and patients with known mental disorder before the diagnosis of PH were excluded. Specific questionnaires were used to assess for depression, anxiety and health related quality of life. Data was collected using electronic medical record including demographics, clinical characteristics, PH class, hemodynamic parameters and treatment detail.

All patients had informed consent for this study, which was approved by the Ethics Committee.

### Assessment of depression, anxiety and health related quality of life

The Hospital Anxiety and Depression Scale (HADS) questionnaire, which was developed in 1983 by Zigmond and Snaith, was used for screening.^12^ The HADS is a fourteen-item scale, seven of the items relate to anxiety and seven to depression. Validity of the HADS was studied by Ingvar Bjelland *et al*., and was found to be a good toll in assessing the symptom severity of anxiety disorders and depression in both somatic, psychiatric and primary care patients and in the general population.^13^ Arabic version of HADS scale has been validated in several studies.^14^ HADS score are from 0-21. Score of 0-7 is considered normal, 8-10 mild or possible, 11-14 moderate or probable and 15-21 severe or definite.^15^ Patients in the category of moderate to severe score were considered to have mental disorder, anxiety and/or depression.

The Short Form 36 (SF-36) Health Survey was used to assess the quality of life in patients with pulmonary hypertension. Health related quality of life (HRQoL) is known as a person’s perceived quality of life representing satisfaction about the areas of life that usually are affected by a chronic illness. HRQoL scores can assess the impact of chronic disease on patient’s quality of life and serve as an indicator of improvement after starting the appropriate management. The SF-36 questionnaire was established in the united states by Boston Health Research Institute. It is mainly used to assess different aspects of the quality of life of adult people. It became the most used method of estimating the health status of the general population as it is easy to use and interpret. The reliability and validity of the SF-36 questionnaire have been assessed in multiple studies worldwide.^16-17^ It involves eight scaled scores, which are the results of the average sum of specific question for each scale. Each scale is graded from 0 to 100, the higher the score the less disability and vice versa.

### Statistical Analysis

Descriptive analysis was used to summarize all variables. Results were reported as mean and standard deviation for continuous variable and categorical variable were calculated as counts and percentages (%). Difference between groups were analyzed using independent t-test (or Man–Whitney test for non-normal data) for continuous variables and chi-square test or Fisher’s exact test for categorical variables. A 5% significance level was employed for all analyses.

### Ethical consideration

The study was conducted in accordance with ethical principles in the Helsinki declaration and the policies and guidelines for clinical research as set forth by office of research affairs at the institution and approved by the ethical review committee. The study was approved by Research Advisory Committee (RAC) of King Faisal Specialist Hospital & Research Center, Riyadh, Saudi Arabia. The RAC is supported by four standing committees; the Research Ethics Committee (REC), the Clinical Research Committee (CRC), the Basic Research Committee (BRC), and the Animal Care and Use Committee (ACUC) and administratively by the Office of Research Affairs (ORA). This study received full approval from RAC including from Research Ethical Committee (REC). The REC, the Institutional Review Board (IRB), evaluates the ethical aspects of all research proposals that involve human subjects. Project Approval was granted with approval ID# 2161034 Informed consent was obtained in view of prospective nature of study and it did not involve any therapeutic intervention.

## Results

Forty-two patients were included in the study with a mean age of 42.8 (±13.3) years and 31(74%) were female. Nineteen (45%) patients were married, 18(43%) were financially independent and 14(33%) had post-secondary education. Mean pulmonary artery pressure (mPAP) of the cohort was 46.4(±15.1) mmHg. Twenty-five (60%) patients had severe, 12(28%) moderate and 5(12%) mild PH (mPAP ≥45, 35-44 and 25-34 mmHg respectively). Seventeen (40%) patients were on single pulmonary vasodilator treatment while the rest on combination therapy. Twelve (30%) patients had co-morbid conditions including diabetes mellitus, hypertension, hypothyroidism and chronic kidney disease. Thirty-eight (90%) patients had World Health Organization Functional Class (WHO-FC) II/III and 4 (10%) class IV (Table 1). None of the patients reported substance abuse.

**Table 1.** Demographic and clinical characteristics

| Characteristics | Value |
| --- | --- |
| Age, mean ±SD | 42.8 ± 13.3 |
| Females, n(%) | 31(74) |
| **WHO Functional Class, n(%)** |  |
| II/III | 38(90) |
| IV | 4(10) |
| **PH Group, n(%)** |  |
| Group 1 | 29(69) |
| Group 4 | 9((21) |
| Other | 4(10) |
| **Severity of PH, n(%)** |  |
| Mild | 5(12) |
| Moderate | 12(28) |
| Severe | 25(60) |
| **HADS Anxiety score, mean ±SD** |  |
| Overall, n=42 | 5.4±4 |
| Normal to mild (0-10), n=38 | 4.5±2.7 |
| Moderate to severe (10-21), n=4 | 14.0±2.9 |
| **HADS Depression score, mean ±SD** |  |
| Overall, n=42 | 4.9±6 |
| Normal to mild (0-10), n=36 | 3.3±2.7 |
| Moderate to severe (10-21), n=6 | 13.8±2.8 |

Mean HADS anxiety and depression score overall was 5.4(±4) and 4.9(±6) respectively. HADS anxiety score was 0-10 (normal-mild) in 38 (90%) and 4 (10%) patients had score of 11-21 (moderate-severe). Thirty-six patients (86%) had HADS depression score 0-10 and 6(14%) had 11-21(Fig 1). There was no significant difference in severity of pulmonary hypertension between normal-mild and moderate-severe HADS score group (*p* 0.219 for anxiety and *p* 0.312 for depression) (Fig 2). Presence of anxiety and/or depression affected five of the components of Quality of life (SF-36) assessment namely physical functioning, physical role performance, vitality, emotional well-being and social functioning (*p* <0.05).

**Fig 1.**
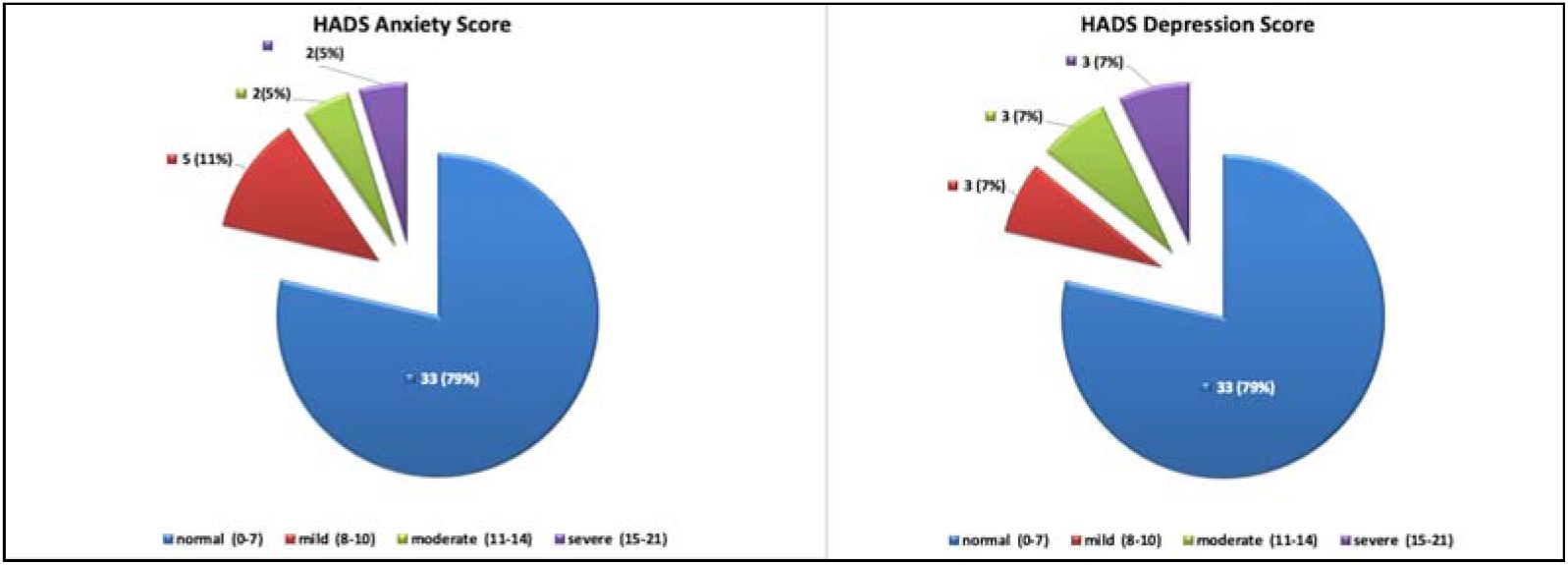
HADS Anxiety and Depression Score

**Fig 2.**
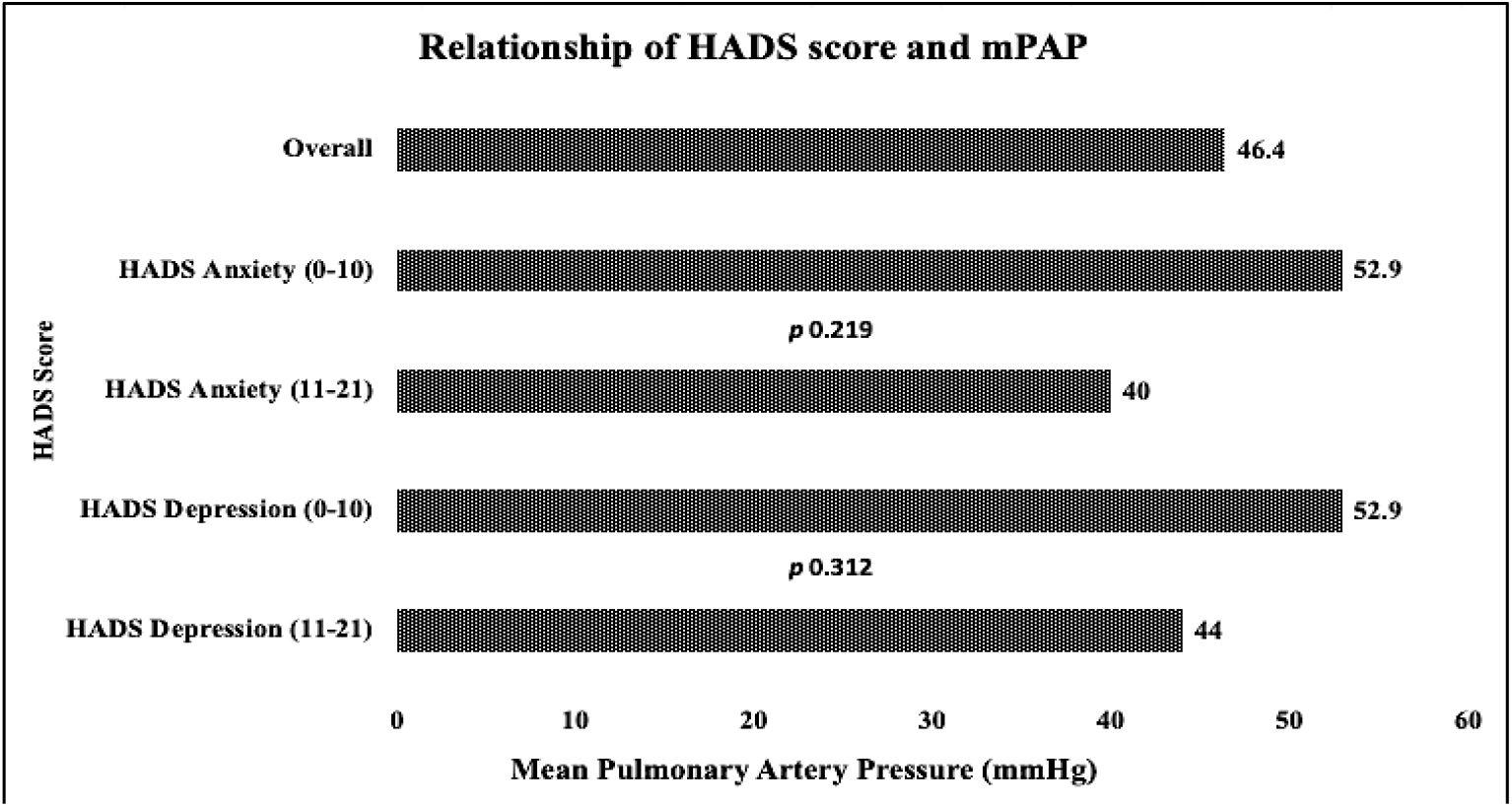
Relationship of HADS score and mean Pulmonary Artery Pressure

There was no significant difference in bodily pain, general health perception and emotional role performance scores (Table 2, Fig 3).

**Table 2.** Pulmonary hemodynamics and correlation of HADS and SF-36 score

| Characteristics | Mean ± SD<br>N=42 | HADS score 0-10<br>(Normal/Mild) |  | HADS score 11-21<br>(Moderate/Severe) |  | P Value |  |
| --- | --- | --- | --- | --- | --- | --- | --- |
|  |  | Anxiety<br>N=38 | Depression<br>N=36 | Anxiety<br>N=4 | Depression<br>N=6 | Anxiety | Depression |
| **Pulmonary hemodynamics** |  |  |  |  |  |  |  |
| Right Atrial Pressure (RAP) | 11.8 ± 5.5 | 11.4 ± 5.5 | 11.9 ± 5.8 | 15.5 ± 4.5 | 11.5 ± 3.7 | 0.1574 | 0.8866 |
| Cardiac Index (CI) | 2.5 ± 0.8 | 2.5 ± 0.8 | 2.5 ± 0.8 | 2.69 ± 0.66 | 2.6 ± 0.4 | 0.6662 | 0.7746 |
| mPAP* | 46.4 ± 15.1 | 52.9 ± 20.3 | 52.9 ± 20.8 | 40 ± 10 | 44 ± 9.4 | 0.2199 | 0.3123 |
| **Quality of life (SF-36)** |  |  |  |  |  |  |  |
| Physical Functioning | 56.5 ± 30.1 | 59.5 ± 30.1 | 61.9 ± 27.8 | 28.8 ± 8.5 | 24.2 ± 23.1 | 0.0508 | 0.0032 |
| Role limitation-physical health | 42.3 ± 45.7 | 46.7 ± 45.8 | 46.5 ± 45.6 | 0 ± 0 | 16.7 ± 40.8 | <.0001 | 0.1403 |
| Role limitation-emotional | 66.7 ± 33.3 | 52.7 ± 47.5 | 51.9 ± 48.8 | 16.6 ± 19.1 | 33.3 ± 29.9 | 0.1438 | 0.3749 |
| Energy/Fatigue | 69.5 ± 28.2 | 60.0 ± 28.4 | 60.8 ± 28.7 | 25.0 ± 17.8 | 31.7 ± 19.7 | 0.0211 | 0.0220 |
| Emotional well being | 57.3 ± 17.8 | 74.7 ± 23.8 | 75.9 ± 22.4 | 20 ± 17.6 | 30.7 ± 8.9 | <.0001 | <.0001 |
| Social Functioning | 70 ± 34.4 | 75.3 ± 30.8 | 75 ± 32.7 | 18.8 ± 23.9 | 39.6 ± 30 | 0.0010 | 0.0175 |
| Pain | 49.2 ± 46.7 | 68.7 ± 32.1 | 69.4 ± 32.1 | 48.1 ± 43.8 | 50.4 ± 38.5 | 0.2448 | 0.1987 |
| General Health | 56.7 ± 29.3 | 58.8 ± 17.7 | 59 ± 18.1 | 42.5 ± 11.9 | 45 ± 30.4 | 0.0807 | 0.0674 |

\*Mean Pulmonary Artery Pressure

**Fig 3.**
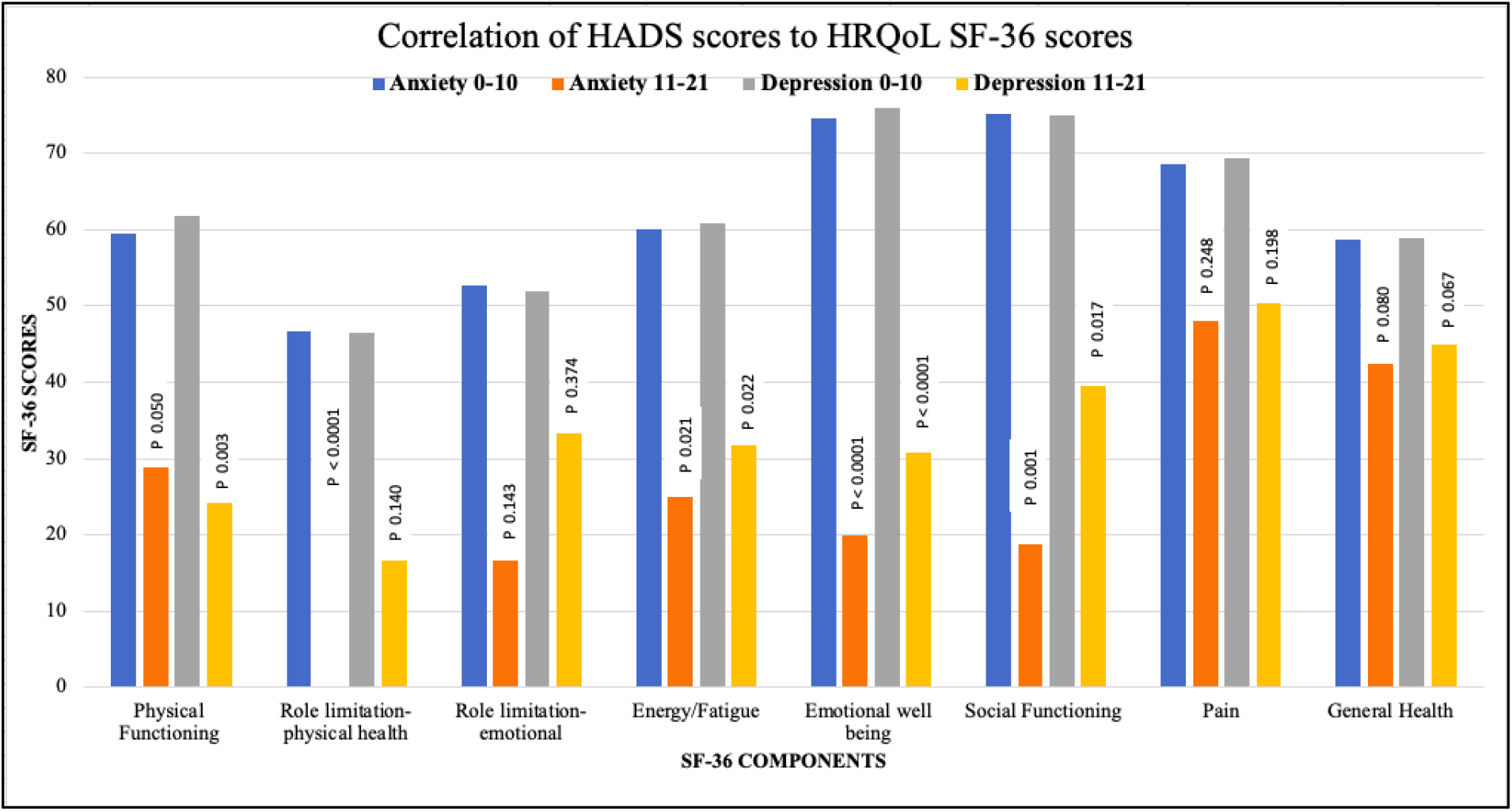
Correlation of HADS scores to HRQoL SF-36 scores

## Discussion

Pulmonary Hypertension is a chronic progressive and debilitating disease. The presence of exertional dyspnea and fatigue, being the most common symptoms,^18^ affect the daily activities and quality of life. Presence of anxiety and/or depression may further deteriorate the already compromised quality of life. The reported prevalence of anxiety and depression in patients with pulmonary hypertension is high and HADS score has been shown to correlate with HRQoL.^19^

This prospective observational study, for the first time, shows the prevalence of anxiety and depression in PH patients in this particular region. The low prevalence of mood disorders (10% anxiety and 14% depression) in this cohort is very different than the published studies where the prevalence is significantly high. This difference can be explained by the social, cultural and family setup in Saudi Arabia where joint family living and close social contact and support play important role in alleviating the patient’s mental suffering. The fact that the presence of mood disorders in this study does not show any correlation to severity of pulmonary hypertension, these disorders may be caused by other factors such as social isolation, unemployment and lack of financial support. The role of family and social support may help to prevent these disorders and improve the quality of life.^20^

## Conclusion

This is the first study highlighting the presence of mood disorders in PH patients in this region. Prevalence of anxiety and depression in PH patients in Saudi Arabia is low (10% and 14% respectively) compared to published studies. The low prevalence may be explained by close family ties, cultural differences and organized social services.

## Limitation of Study

The main limitation of study is small size of the cohort. Larger studies are needed to assess the true prevalence of anxiety and depression in PH patients in this region.

## Data Availability

Data is available

## Abbreviations

PH: Pulmonary Hypertension
HADS: Hospital Anxiety and Depression Scale
HRQoL: Health Related Quality of Life
SF-36: Short Form 36
FC: Functional Class
mPAP: mean Pulmonary Arterial Pressure
PVR: Pulmonary Vascular Resistance
PCWP: Pulmonary Capillary Wedge Pressure

## Reference

1. Kiely DG, Elliot CA, broe I, Condliffe R. Pulmonary hypertension: diagnosis and management. BMJ. 2013;346:f2028. Published 2013 Apr 16. doi:10.1136/bmj.f2028

2. Badesch DB, Raskob GE, Elliott CG, et al. Pulmonary arterial hypertension: baseline characteristics from the REVEAL Registry. Chest. 2010;137(2):376–387. doi:10.1378/chest.09-1140

3. Flattery MP, Pinson JM, Savage L, Salyer J. Living with pulmonary artery hypertension: patients’ experiences. Heart Lung. 2005;34(2):99–107. doi:10.1016/j.hrtlng.2004.06.010

4. Guillevin L, Armstrong I, Aldrighetti R, et al. Understanding the impact of pulmonary arterial hypertension on patients’ and carers’ lives. Eur Respir Rev. 2013;22(130):535–542. doi:10.1183/09059180.00005713

5. Löwe B, Gräfe K, Ufer C, et al. Anxiety and depression in patients with pulmonary hypertension. Psychosom Med. 2004;66(6):831–836. doi:10.1097/01.psy.0000145593.37594.39

6. McCollister DH, Beutz M, McLaughlin V, et al. Depressive symptoms in pulmonary arterial hypertension: prevalence and association with functional status. Psychosomatics. 2010;51(4):339-339.e8. doi:10.1176/appi.psy.51.4.339

7. Somaini G, Hasler ED, Saxer S, et al. Prevalence of Anxiety and Depression in Pulmonary Hypertension and Changes during Therapy. Respiration. 2016;91(5):359–366. doi:10.1159/000445805

8. Badesch DB, Raskob GE, Elliott CG, et al. Pulmonary arterial hypertension: baseline characteristics from the REVEAL Registry. Chest. 2010;137(2):376–387. doi:10.1378/chest.09-1140

9. Guillevin L, Armstrong I, Aldrighetti R, et al. Understanding the impact of pulmonary arterial hypertension on patients’ and carers’ lives. Eur Respir Rev. 2013;22(130):535–542. doi:10.1183/09059180.00005713

10. Galiè N, Humbert M, Vachiery JL, et al. 2015 ESC/ERS Guidelines for the diagnosis and treatment of pulmonary hypertension: The Joint Task Force for the Diagnosis and Treatment of Pulmonary Hypertension of the European Society of Cardiology (ESC) and the European Respiratory Society (ERS): Endorsed by: Association for European Paediatric and Congenital Cardiology (AEPC), International Society for Heart and Lung Transplantation (ISHLT) [Published correction appears in Eur Respir J. 2015 Dec;46(6):1855-6]. Eur Respir J. 2015;46(4):903–975. doi:10.1183/13993003.01032-2015

11. Idrees MM, Saleemi S, Azem MA, et al. Saudi guidelines on the diagnosis and treatment of pulmonary hypertension: 2014 updates. Ann Thorac Med. 2014;9 (Suppl 1):S1-S15. doi:10.4103/1817-1737.134006

12. Zigmond AS, Snaith RP. The hospital anxiety and depression scale. Acta Psychiatr Scand. 1983;67(6):361–370. doi:10.1111/j.1600-0447.1983.tb09716.x

13. Bjelland I, Dahl AA, Haug TT, Neckelmann D. The validity of the Hospital Anxiety and Depression Scale. An updated literature review. J Psychosom Res. 2002;52(2):69–77. doi:10.1016/s0022-3999(01)00296-3

14. Terkawi AS, Tsang S, AlKahtani GJ, et al. Development and validation of Arabic version of the Hospital Anxiety and Depression Scale. Saudi J Anaesth. 2017;11(Suppl 1):S11–S18. doi:10.4103/sja.SJA_43_17

15. Anna F. Stern, The Hospital Anxiety and Depression Scale, Occupational Medicine, Volume 64, Issue 5, July 2014, Pages 393–394, https://doi.org/10.1093/occmed/kqu024

16. Lins L, Carvalho FM. SF-36 total score as a single measure of health-related quality of life: Scoping review. SAGE Open Med. 2016;4:2050312116671725. Published 2016 Oct 4. doi:10.1177/2050312116671725

17. Brazier JE, Harper R, Jones NM, et al. Validating the SF-36 health survey questionnaire: new outcome measure for primary care. BMJ. 1992;305(6846):160–164. doi:10.1136/bmj.305.6846.160

18. Bazan IS, Fares WH. Pulmonary hypertension: diagnostic and therapeutic challenges. Ther Clin Risk Manag. 2015;11:1221–1233. Published 2015 Aug 17. doi:10.2147/TCRM.S74881

19. Harzheim D, Klose H, Pinado FP, et al. Anxiety and depression disorders in patients with pulmonary arterial hypertension and chronic thromboembolic pulmonary hypertension. Respir Res. 2013;14(1):104. Published 2013 Oct 9. doi:10.1186/1465-9921-14-104

20. Hwang B, Howie-Esquivel J, Fleischmann KE, Stotts NA, Dracup K. Family caregiving in pulmonary arterial hypertension. Heart Lung. 2012;41(1):26–34. doi:10.1016/j.hrtlng.2011.03.002

